# Mapping Global Research Trends in Digital Occupational Health: A Bibliometric Analysis Utilizing the Scopus Database

**DOI:** 10.1101/2024.04.18.24306040

**Authors:** Youssef Er-Rays, Meriem M’dioud, Hamid Ait Lemqeddem

## Abstract

The 2019 health crisis has underscored the imperative role of information and communications technology (ICT) in bolstering employees’ health, well-being, and ameliorating health risks associated with artificial intelligence (AI) errors. Digital health technology facilitates remote healthcare delivery, employee health monitoring, and enhanced communication channels between workers and healthcare professionals. This paper conducts a comprehensive analysis of global trends in digital occupational health research over the past decade, with a specific focus on prolific authors, countries, academic institutions, and publications. Leveraging the PRISMA framework and VOSviewer software, relevant documents are extracted from the Scopus database. The findings underscore the significance of digital technology in the realm of occupational health and highlight the pressing need for further investigation, particularly encompassing emerging economies and employing robust methodologies to ensure the reliability of results. This study offers a valuable assessment of advancements in digital occupational health, serving as a foundation for future research endeavors aimed at optimizing workplace health and well-being on a global scale.

## 1. Introduction

The COVID-19 pandemic has underscored the pivotal importance of digital technology in advancing workplace health initiatives and bolstering employee well-being. Various digital health technologies, such as remote healthcare delivery, employee well-being monitoring systems, seamless communication channels between employees and healthcare providers, and real-time provision of health and safety information, have experienced widespread adoption. Additionally, health and wellness applications have played a crucial role in helping employees maintain healthy lifestyles. For instance, Blake, Somerset, and Greaves (2022) developed the Pain at Work (PAW) toolkit, designed to aid employees in managing chronic or persistent pain while at work. Similarly, Test@Work, introduced and evaluated by Blake, Somerset, and Evans (2020), aimed to enhance employers’ understanding of workplace health screenings and HIV testing procedures. Furthermore, Blake, Lai, Coman, Houdmont, and Griffiths (2019) conducted an assessment of a digital intervention targeting sedentary office employees in China, revealing positive outcomes associated with the adoption of digital tools.

These contributions highlight the pivotal role that digital technology plays in advancing workplace health and well-being initiatives. They showcase how digital tools can empower employees to better manage their health conditions, improve employers’ understanding of health-related issues in the workplace, and promote healthier lifestyles among sedentary office workers. As organizations continue to navigate the challenges posed by the pandemic and prioritize employee health and well-being, the adoption and utilization of digital health technologies are likely to remain integral components of workplace health strategies.

Despite significant expenditures and governments’ backing, a systematic analysis undertaken by Black et al. (2011) finds a lack of empirical data to back up the claimed benefits of digital occupational health solutions. However, Judson et al. (2020) demonstrated the effectiveness of digital chatbots in screening healthcare workers for COVID-19 symptoms, whereas Vehko et al. (2019) highlighted the time pressures and psychological distress experienced by nurses as a result of the integration of electronic health records. As a result, there is an urgent need to expand digital health systems and increase healthcare workers’ expertise in using these technologies to improve patient care. These results highlight the vital need of taking an evidence-based approach to the deployment of digital health technology, with an emphasis on user experience.

Given the results reported by Black et al. (2011), which emphasize the lack of empirical validation for the advantages of digital occupational health tools, it is clear that further study is required to determine their usefulness and impact. While Judson et al. (2020) demonstrated a successful use of digital chatbots in healthcare professional screening, and Vehko et al. (2019) provided light on the issues that nurses confront, it is critical to increase this body of data via careful inquiry and analysis. This involves investigating the usability, usefulness, and possible downsides of various digital health solutions in different healthcare settings and demographics.

Moving ahead, healthcare organizations and governments must emphasize the development and implementation of evidence-based approaches to incorporating digital health technology into clinical practice (Er Rays, Lequaddem, et Ezzahiri 2024; Er-Rays et al. 2024a, 2024b; Er-Rays et Ait-Lemqeddem 2020; Er-Rays et Lemqeddem 2020, 2021; Er-Rays et M’dioud 2024a, 2024b). This includes investing in R&D activities that prioritize user-centered design, interoperability, data security, and regulatory compliance. Furthermore, continuing education and training programs should be implemented to ensure that healthcare personnel have the essential skills and expertise to properly use these technologies in their everyday practice. By taking a comprehensive strategy that considers both technical and human elements, the healthcare sector can maximize the advantages of digital health solutions while reducing possible dangers and obstacles.

The main objective of this research is to conduct an extensive examination of worldwide developments in digital occupational health research over the past decade. Its goal is to identify influential authors, countries, academic institutions, and journals, quantifying pertinent articles and citations relevant to marketing studies. Moreover, the study delves into network overlay visualization of keywords among references. Diverging from other review papers in neuromarketing, this research places emphasis on elucidating global trends in digital occupational health research, striving for a precise, comprehensive, and succinct conclusion. The primary contributions of this bibliometric survey entail a thorough analysis of selected documents, encompassing annual and cumulative publications, as well as productive journals from 2010 to 15/02/2024. Additionally, it offers comprehensive insights into recent global trends in digital/occupational health, introducing new references and providing guidance for emerging researchers in the field.

This work’s structure consists of four essential parts. First, it describes the materials and procedures used. Second, it presents the findings from the bibliometric study. Third, it discusses the consequences of these discoveries. Finally, it addresses the study’s shortcomings and makes suggestions for future research initiatives.

## 2 Methods

The authors conducted a bibliometric analysis to investigate evolving trends in digital occupational health research. They systematically retrieved pertinent documents from the Scopus database, adhering to the PRISMA framework for a structured approach. Following Block & Fisch’s (2020) guidelines, they meticulously conducted a comprehensive bibliometric analysis, illuminating the field’s organization by identifying active countries, academic institutions, journals, and authors.

Utilizing VOSviewer software facilitated the creation of a visual representation of the scholarly landscape, as illustrated in Figure 1. These methodologies offer valuable insights into the progression of digital occupational health, rendering this study a pivotal reference for scholars delving deeper into this field. Importantly, the authors cited previous instances where VOSviewer proved effective across diverse bibliometric investigations, including digital occupational health.

**Fig. 1.**
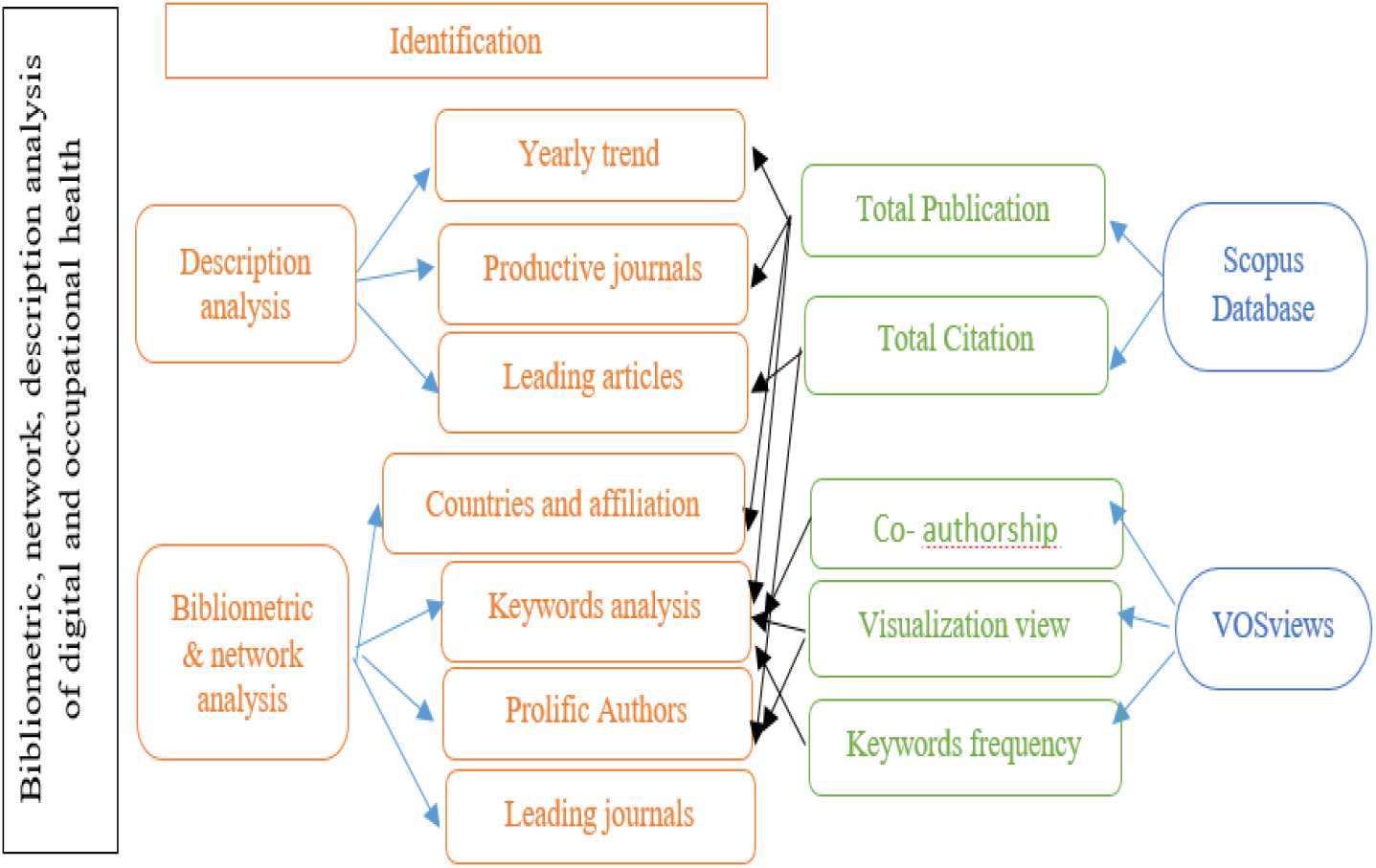
Analytical structure of the current paper

**Fig. 2.**
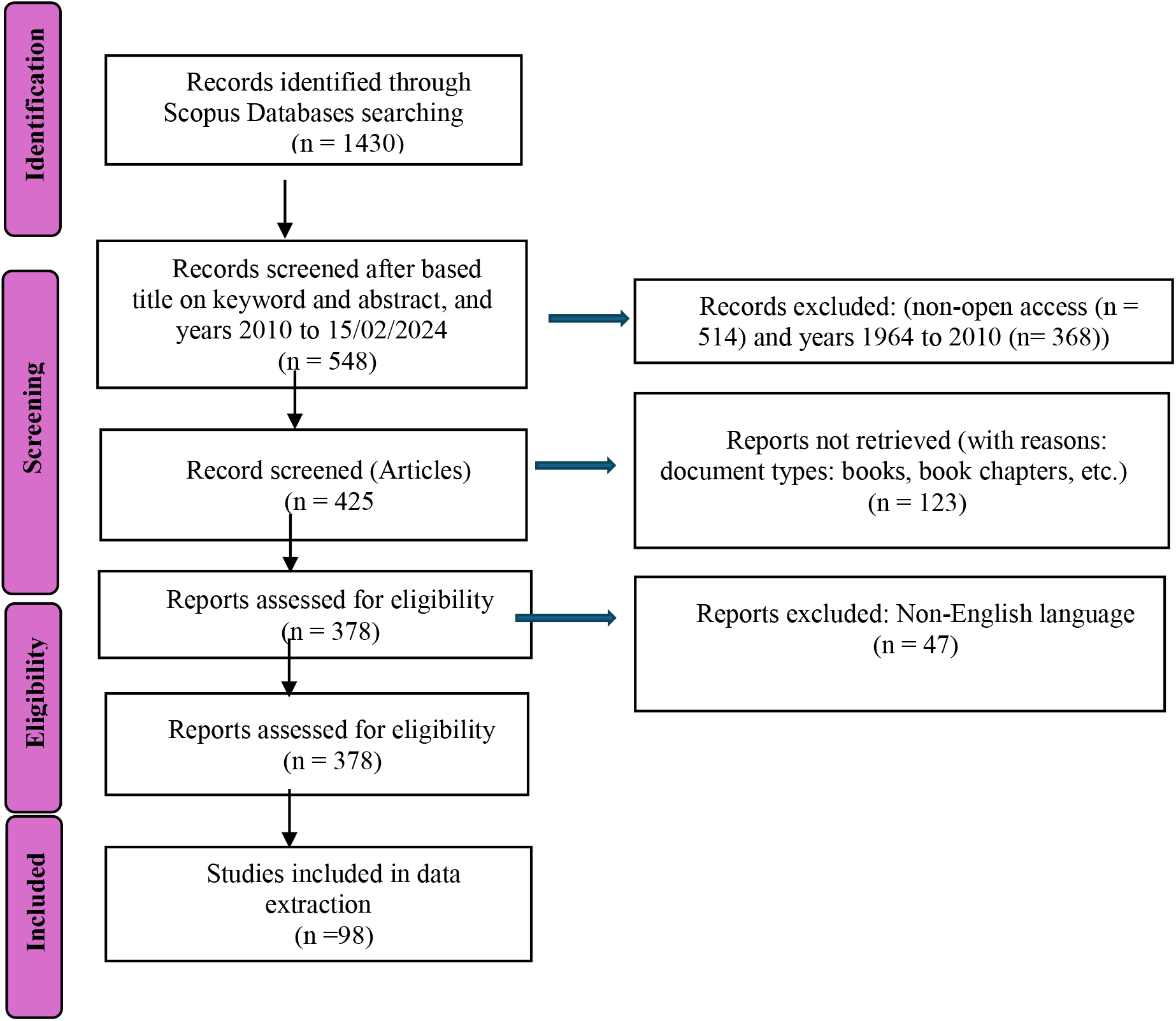
PRISMA flow chart for selecting documents for this study.

### 2.1 Search Criteria

In February 2024, data collecting began using the Scopus database, a well-known key resource in academic circles for finding relevant material within certain study topics. Using a search query that included the phrases (TITLE-ABS-KEY (digital AND occupational AND health), the researcher selected publications published between 2010 and February 2024. This procedure produced a total of 378 documents, 98 of which were carefully selected for in-depth investigation. The selection process followed the PRISMA framework (Moher et al., 2015), which consisted of four key stages: initial identification of relevant publications, subsequent screening, rigorous evaluation of eligibility criteria, and final selection and inclusion of studies that met predefined research criteria.

Throughout the selection process, we took great effort to include research that directly addressed the nexus of digital technology and occupational health. Each found document was rigorously evaluated for its relevance to the study topic, paying special emphasis to methodological rigor, empirical evidence, and contribution to the wider debate on digital occupational health. Studies that matched these criteria were included in the final analysis, whereas those judged irrelevant or lacking methodological robustness were omitted. This rigorous process guaranteed that high-quality papers were chosen that may provide useful insights into trends and changes in digital occupational health research across the given time period.

This paper examines worldwide digital occupational health research trends over the last decade, focusing on prominent authors, countries, academic institutions, and publications. The research used the PRISMA framework (Moher & al., 2015) to discover 1,430 relevant publications using the query “TITLE-ABS-KEY (digital AND occupational AND health).” The dataset was then filtered, which reduced it to 425 documents. Eligibility was established, resulting in the inclusion of 378 papers that met certain requirements. The research then chose and included 98 relevant publications to ensure the academic quality and robustness of the investigation. The thorough selection procedure assures the study’s strength and quality.

The study’s scope was meticulously delineated to ensure relevance and precision. Documents such as articles and reviews specifically focusing on digital occupational health from 2010 to 2024 were incorporated to capture the field’s comprehensive evolution over the decade. Additionally, documents leveraging digital tools within occupational health were intentionally included, recognizing their significant influence on shaping research in this domain. To maintain consistency, documents published in languages other than English and those classified as book chapters or conference materials were excluded, aligning the study’s focus exclusively with scholarly articles and reviews to uphold academic rigor. This rigorous selection process was instrumental in facilitating a robust and meaningful analysis of trends in digital occupational health.

### 2.2 Materials and Methods

Utilizing bibliometric analysis, the researcher meticulously scrutinized emerging trends in digital occupational health research. The study’s structured effectiveness was ensured through the application of the PRISMA framework, facilitating the systematic retrieval of relevant documents from the Scopus database. Following guidelines established by Block & Fisch (2020), the author executed a compelling bibliometric analysis, unveiling key insights into the field’s landscape, including prominent countries, academic institutions, journals, and authors. Figure 1 visually represents this scholarly landscape, crafted using VOSviewer software. These methodical processes offer valuable insights into the evolution of digital occupational health, serving as a vital resource for scholars initiating further research ventures.

While specific authorship is not explicitly mentioned, data collection occurred in April 2023. Employing a comprehensive query, (TITLE-ABS-KEY (digital AND occupational AND health)), publications from 2010 to February 2024 were sought, yielding 378 documents. Adhering to the PRISMA framework’s rigorous four-stage methodology—identification, screening, eligibility assessment, and selection—98 papers were meticulously chosen for thorough analysis. This meticulous approach ensures the robustness and scholarly integrity of the study’s findings.

## 3 Results

### 3.1 Descriptive Analysis

#### Growth of the Publication

Figure 3 presents the annual publication trends in digital occupational health research based on data retrieved from the Scopus database. The table illustrated the number of publications per year from 2013 to 2023, as well as the cumulative publication count over the same period.

**Fig. 3.**
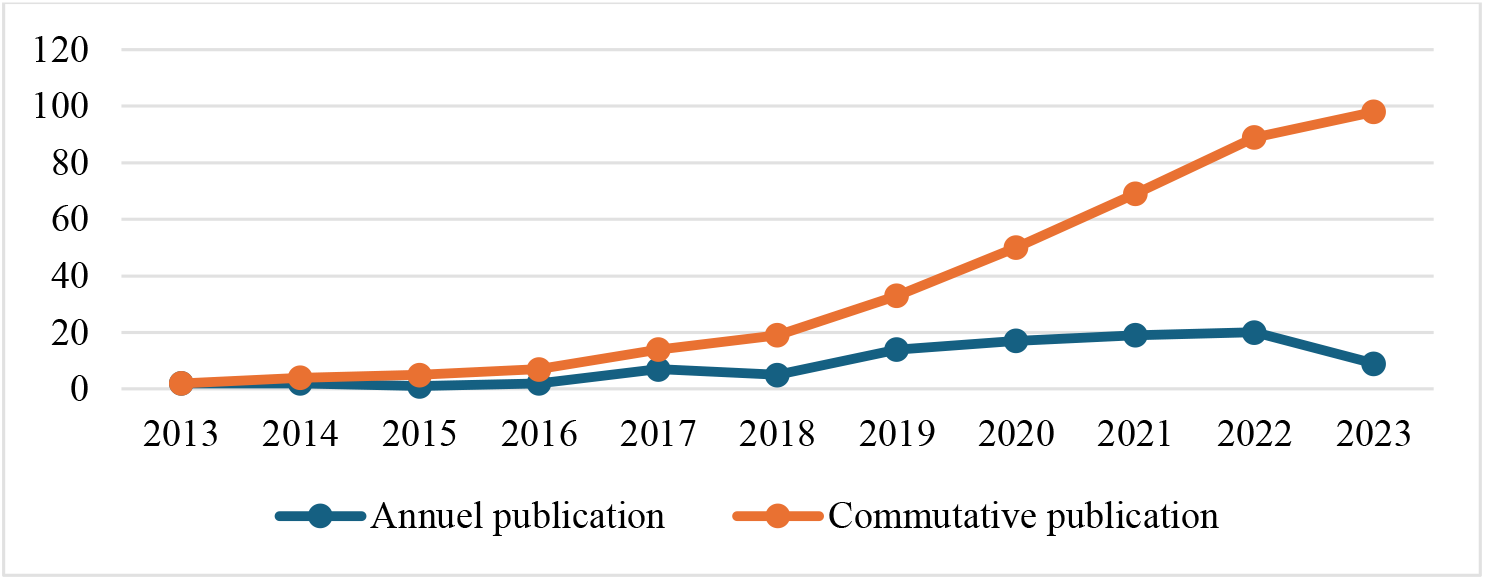
Annual publication on digital occupational health using Scopus database

**Fig. 4.**
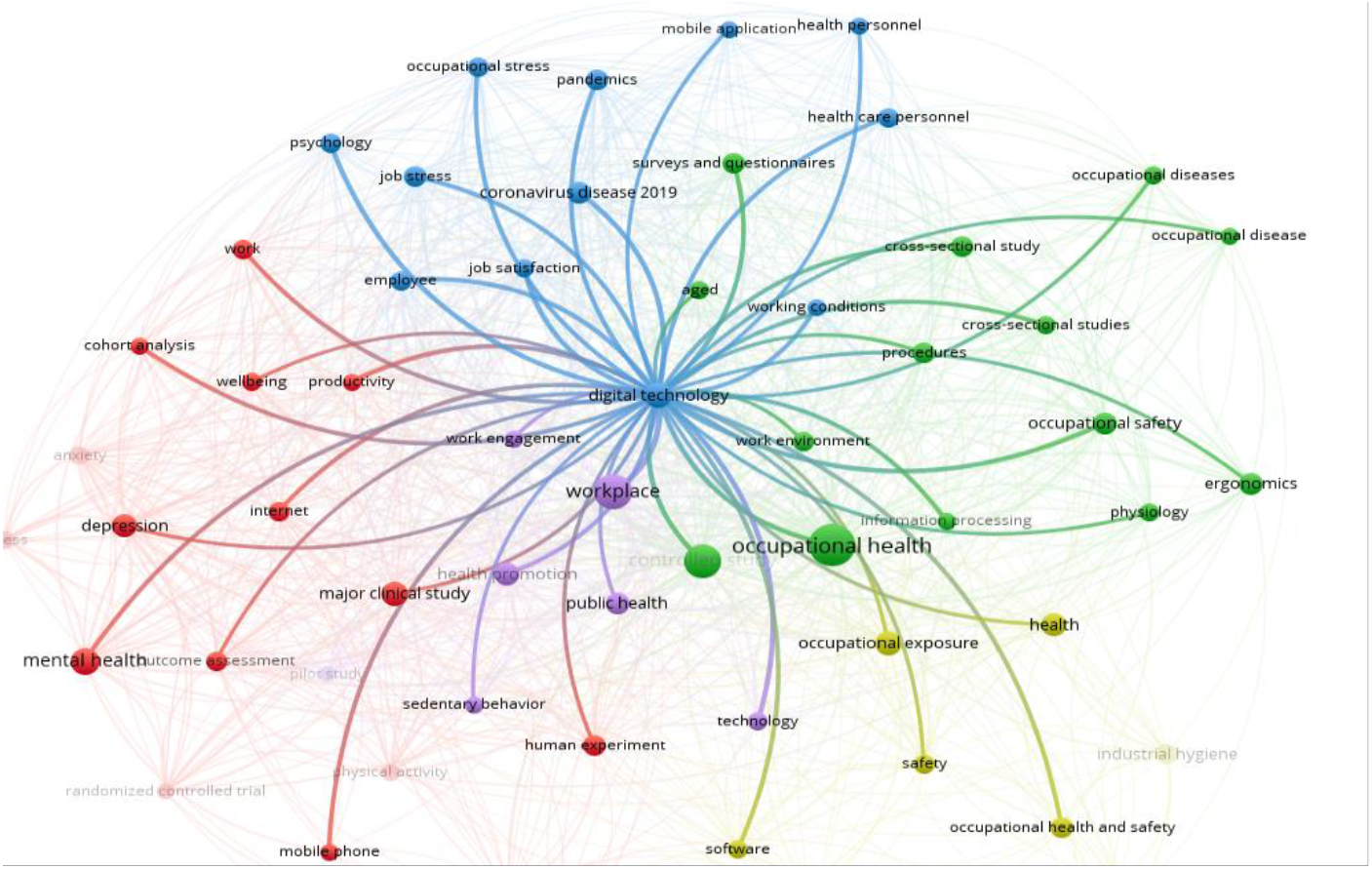
Network visualization for 69 keywords co-occurrence (with min. 5 occurrence)

The annual publication count shows variations across the years. From 2013 to 2016, the number of publications remained relatively low, ranging from 1 to 2 publications annually. However, there was a notable increase in 2017, with 7 publications recorded, followed by fluctuations in subsequent years. The peak in annual publications occurred in 2021, with 19 publications, indicating a significant surge in research activity during that year. However, there was a slight decline in 2022 and a further decrease in 2023, with 20 and 9 publications, respectively.

The cumulative publication count, on the other hand, demonstrates a consistent upward trajectory. Starting from 2 publications in 2013, the cumulative count steadily increases over the years, reaching 98 publications by the end of 2023. This indicates a cumulative growth in the body of literature on digital occupational health research over the specified period.

#### Journal Outlets

Table 1 provides an overview of the most productive journals in the field of digital and occupational health, considering those with a minimum of 2 documents. The table includes the total production (TP) of each journal, the publisher, country of origin, reference of the most cited document, and the number of times cited.

**Table 1.**
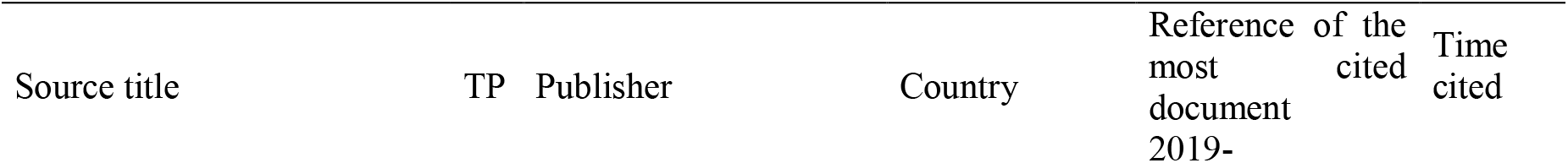
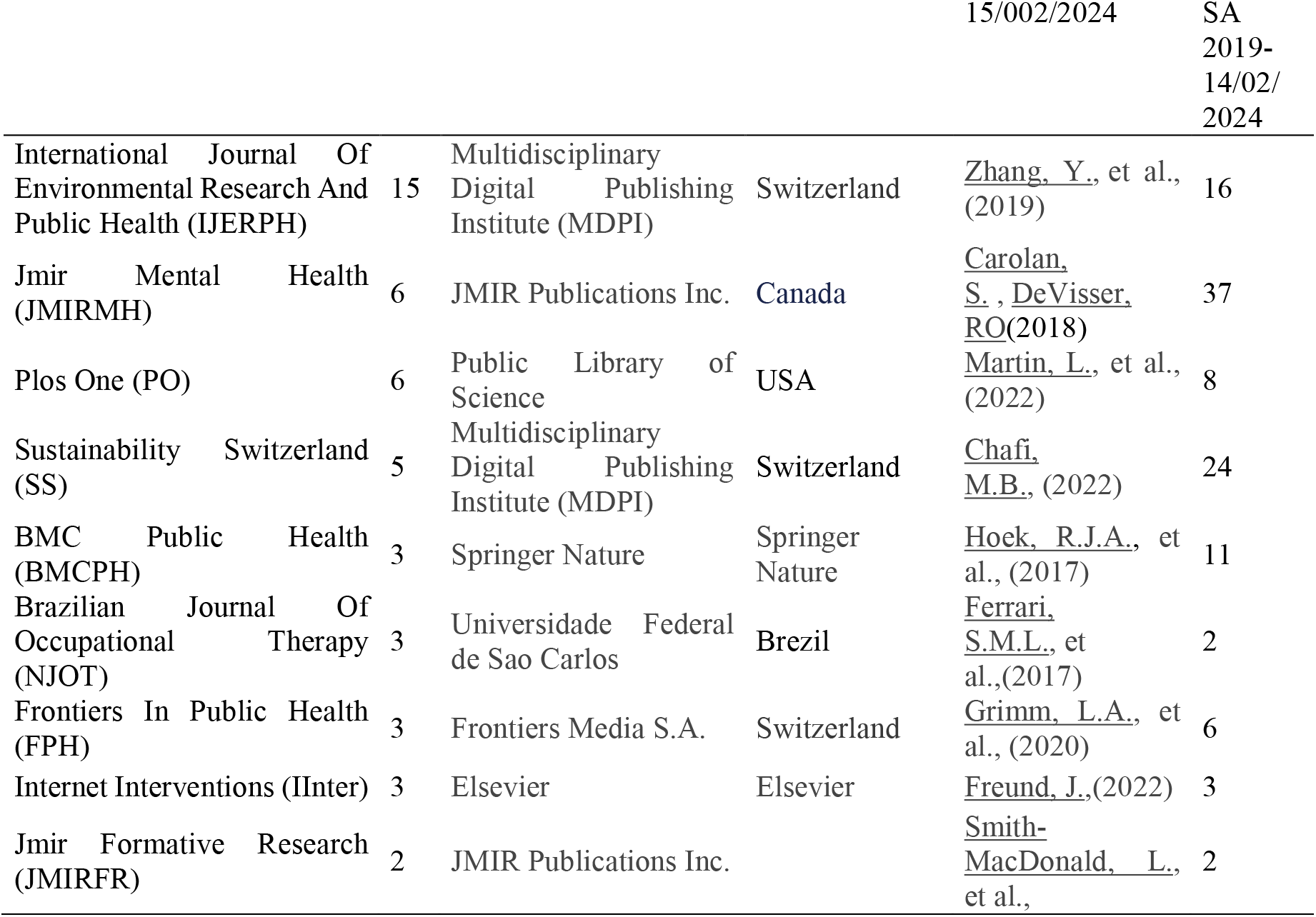
The most Productive journal in digital and occupational health (with min. 2 documents).

| Source title | TP | Publisher | Country | Reference of the most cited document | Time cited |
| --- | --- | --- | --- | --- | --- |
|  |  |  |  | 2019- |  |
| International Journal Of Environmental Research And Public Health (IJERPH) | 15 | Multidisciplinary Digital Publishing Institute (MDPI) | Switzerland | <u>Zhang, Y.</u>, et al., (2019) | 16 |
| Jmir Mental Health (JMIRMH) | 6 | JMIR Publications Inc. | Canada | <u>Carolan, S.</u>, <u>DeVisser, R.O.</u>(2018) | 37 |
| Plos One (PO) | 6 | Public Library of Science | USA | <u>Martin, L.</u>, et al., (2022) | 8 |
| Sustainability (SS) | 5 | Multidisciplinary Digital Publishing Institute (MDPI) | Switzerland | <u>Chafi, M.B.</u>, (2022) | 24 |
| BMC Public Health (BMCPH) | 3 | Springer Nature | Springer Nature | <u>Hoek, R.J.A.</u>, et al., (2017) | 11 |
| Brazilian Journal Of Occupational Therapy (NJOT) | 3 | Universidade Federal de Sao Carlos | Brazil | <u>Ferrari, S.M.L.</u>, et al.,(2017) | 2 |
| Frontiers In Public Health (FPH) | 3 | Frontiers Media S.A. | Switzerland | <u>Grimm, L.A.</u>, et al., (2020) | 6 |
| Internet Interventions (IInter) | 3 | Elsevier | Elsevier | <u>Freund, J.</u>,(2022) | 3 |
| Jmir Formative Research (JMIRFR) | 2 | JMIR Publications Inc. |  | <u>Smith-MacDonald, L.</u>, et al., | 2 |

The IJERPH emerges as the most productive journal with a TP of 15. Published by the MDPI in Switzerland, it boasts a highly cited document by Zhang et al. (2019), cited 16 times as of February 14, 2024.

Following closely is JMIRMH, with a TP of 6, published by JMIR Publications Inc. in Canada. A study by Carolan and DeVisser (2018) stands out as the most cited document, cited 37 times.

PO a publication of the Public Library of Science in the USA, also demonstrates significant productivity with a TP of 6. A paper authored by Martin et al. (2022) has garnered 8 citations.

Other notable journals include SS (TP: 5), BMCPH (TP: 3), BJOT (TP: 3), Frontiers in Public Health (TP: 3), IInter (TP: 3), and JMIRFR (TP: 2). These journals represent a diverse range of publishers and countries of origin, indicating the global scope and interdisciplinary nature of research in digital and occupational health.

Overall, the findings from Table 1 highlight the key journals contributing to the scholarly discourse in this field and the impact of their publications as reflected in citation counts. These journals serve as vital platforms for disseminating research findings and fostering collaboration among researchers worldwide.

#### Productive Authors, Countries, and Institutions

Between 2010 and February 15th, 2023, the scholarly landscape witnessed the involvement of 171 authors in digital occupational health research. Notably, Blake, H. and colleagues (2019, 2020, 2022, and 2023) emerged as frontrunners, securing the top position with 26 citations garnered from their collective publication of 5 articles. Following closely, Halldin, C. N., Laney, A. S., Carolan, S., and De Visser, R. O., alongside Carolan, S. and their respective research teams, attained the second position, with each group accruing 33 citations from the dissemination of 3 articles each.

Geographically, the United States emerged as the predominant contributor, with 20 published articles, closely trailed by the United Kingdom with 18 contributions. The Netherlands and Sweden shared the third position, each producing 7 articles, reflecting a balanced distribution of scholarly output among these nations.

Analyzing the institutional landscape reveals the participation of 160 institutions, showcasing a diverse network of contributors. Notably, the University of Nottingham, the University of St. Gallen, and the Nottingham Biomedical Research Centre demonstrated notable involvement, each contributing 4 publications. Additionally, the University of Lausanne (UNIL), the Friedrich-Alexander-Universität Erlangen-Nürnberg, the National Institute for Occupational Safety and Health, and the Universidad de Oviedo each made significant contributions, adding 3 articles to the scholarly discourse. These insights underscore the multifaceted contributions from authors, countries, and institutions to the evolving field of digital occupational health research. landscape.

#### Subject area

Table 2 presents the distribution of document subject areas within the domains of “digital” and “occupational health” across two distinct periods: 2011-2018 and 2019-2022.

**Table 2.**
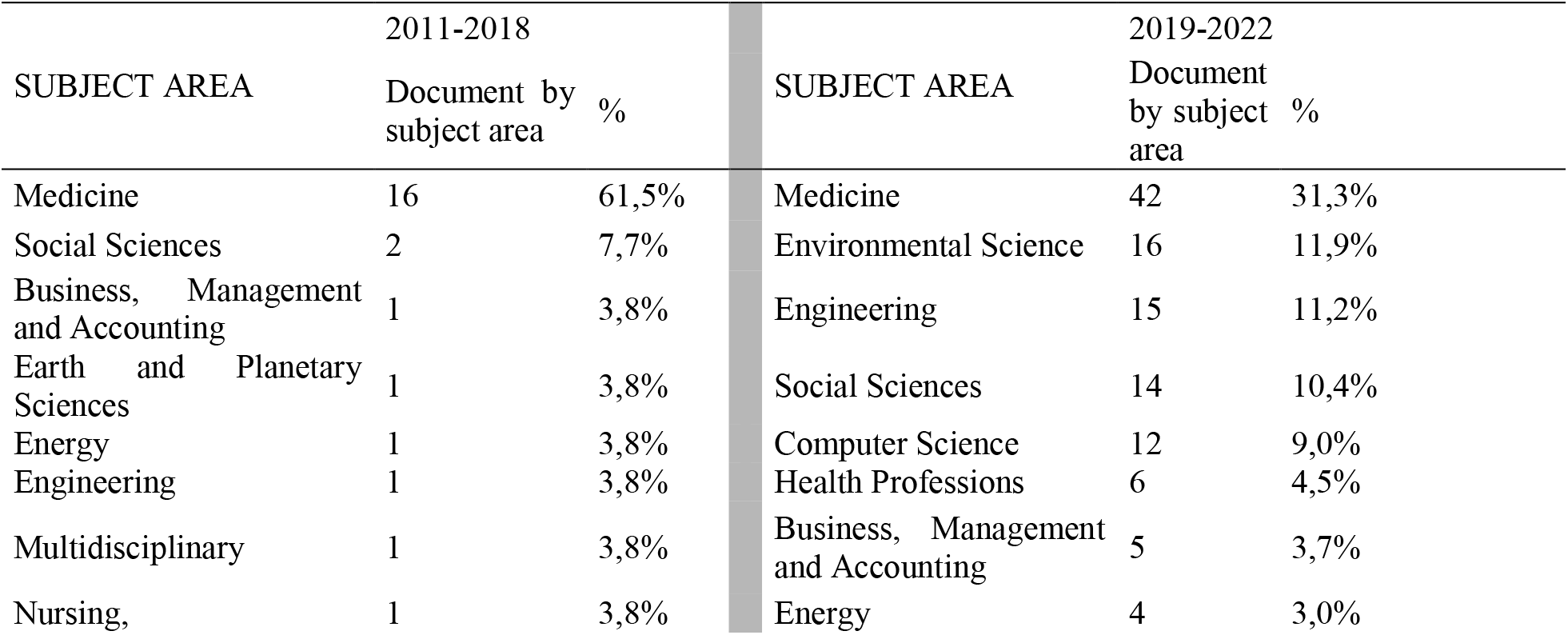
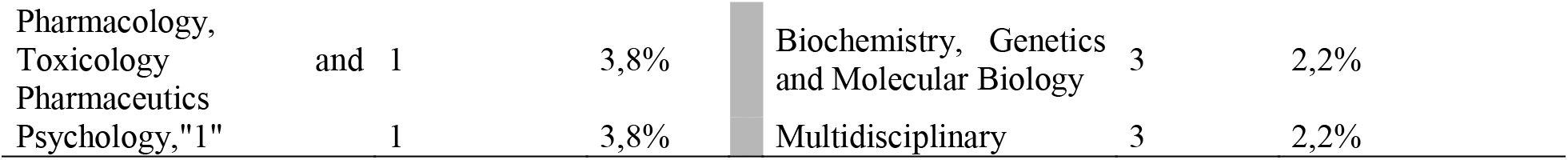
subject area of “digital” and “occupational health,” during two periods 2011-2018 et 2019-2022.

During the period of 2011-2018, the majority of documents were categorized under Medicine, constituting 61.5% of the total, followed by smaller percentages in Social Sciences, Business, Management, and Accounting, Earth and Planetary Sciences, Energy, Engineering, Multidisciplinary, Nursing, Pharmacology, Toxicology, and Pharmaceutics, and Psychology, each accounting for 3.8% to 7.7%.

Conversely, in the subsequent period of 2019-2022, there was a noticeable shift in subject areas. While Medicine remained prominent, its proportion decreased to 31.3%, indicating a relative decline compared to the previous period. Environmental Science emerged as a significant subject area, comprising 11.9% of the documents, followed closely by Engineering and Social Sciences with 11.2% and 10.4% respectively. Computer Science, Health Professions, Business, Management, and Accounting, Energy, Biochemistry, Genetics, and Molecular Biology, and Multidisciplinary subjects constituted smaller percentages ranging from 2.2% to 9.0%.

This transformation underscores evolving research priorities and interdisciplinary collaborations within the domains of digital and occupational health, reflecting a broader engagement across various fields and disciplines.

### 3.2 Bibliometric Analysis

#### Leading Journals

The methodological approach involved identifying the top ten influential journals within the realms of digital and occupational health. “FIJ of Environmental Research and Public Health” emerged as the most prolific among these journals. Additionally, the study assessed the impact of publications using the ACI score, with “JMIRMH” exhibiting the highest score of 23 alongside 92 citations. Notably, a study conducted by Carolan and De Visser (2018) was examined, which investigated employee perspectives on digital mental health interventions within workplace settings, elucidating both obstacles and facilitators to implementation. Importantly, the research underscored the nuanced relationship between publication volume and citation frequency, demonstrating that a journal’s extensive publication output does not inherently guarantee a correspondingly high citation count.

Table 3 provides insights into the most productive journals within the field of digital and occupational health, considering those with a minimum of 2 documents published.

**Table 3.** The most Productive journal in digital and occupational health (with min. 2 documents).

| Source title | TP | Total Citation | ACi |
| --- | --- | --- | --- |
| International Journal Of Environmental Research And Public Health | 12 | 62 | 7 |
| Jmir Mental Health | 5 | 93 | 25 |
| Plos One | 5 | 51 | 14 |
| Sustainability Switzerland | 5 | 42 | 12 |
| BMC Public Health | 3 | 18 | 9 |
| Brazilian Journal Of Occupational Therapy | 3 | 3 | 2 |
| Frontiers In Public Health | 3 | 12 | 7 |
| Internet Interventions | 3 | 29 | 15 |
| Jmir Formative Research | 2 | 5 | 3 |

ACi; average citation per item

The IJERPH emerges as the most prolific journal, with a total production (TP) of 12 documents. These publications have garnered a commendable total citation count of 62, resulting in an average citation per item (ACi) of 7.

Following closely, JMIRMH and PO each contribute 5 documents to the field. While JMIRMH records a substantial total citation count of 93, resulting in an impressive ACi of 25, PO registers 51 citations with an ACi of 14.

Similarly, SS presents 5 documents, accumulating a total citation count of 42 and an ACi of 12, indicating significant recognition within the academic community.

BMCPH, BJOT, FPH, and IInter each contribute 3 documents to the field. BMCPH records a total citation count of 18, resulting in an ACi of 9. The BJOT, on the other hand, registers a lower total citation count of 3, with an ACi of 2. FPH accumulates 12 citations, yielding an ACi of 7, while IInter records 29 citations, resulting in an ACi of 15.

Lastly, JMIRFR adds 2 documents to the body of literature, with a total citation count of 5 and an ACi of 3.

These findings underscore the diversity of journals contributing to the digital and occupational health domain, highlighting their varying levels of impact and recognition within the scholarly community.

#### Keywords Analysis

This study used bibliometric analysis to investigate the interconnection of keywords and their correlations across 98 papers extracted from 61 journals, with a focus on the topics of “digital” and “occupational health” (Wang, M., & Chai, L. (2018)). The intensity of linkage found between these keywords indicates their frequency of occurrence together inside the same articles, with a predetermined threshold of five occurrences necessary for author keywords to appear on the bibliometric map. Using VOSviewer (Van Eck, N., & Waltman, L. (2013)), the study focused on the co-occurrence patterns of 70 terms that fulfilled the minimum occurrence threshold. This technique provides useful insights into the thematic landscape and developing trends within the realm of digital and occupational health research, allowing for a comprehensive knowledge of the key topics and their relationships within the scholarly literature.

Throughout this study, our expectations centered on identifying a robust correlation between the domains of “digital” and “occupational health.” Remarkably, during the COVID-19 crises, we observed the emergence of new thematic clusters (depicted in blue color) related to psychology, well-being, mobility, and the pandemic. Additionally, the analysis highlighted the strong connection between the keyword “digital” and the field of occupational health, encapsulated within the green cluster.

The results presented in Table 4 showcase the top keywords identified through the bibliometric analysis, each with a minimum occurrence of five instances within the dataset. These keywords offer valuable insights into the prevailing trends and focal points within the realm of digital health and occupational health research.

**Table 4.** Top keywords by the minimum 5 occurrences.

| id | keyword | occurrences | total link strength |
| --- | --- | --- | --- |
| 241 | coronavirus disease 2019 | 10 | 151 |
| 277 | depression | 11 | 134 |
| 316 | digital technology | 13 | 148 |
| 366 | employee | 8 | 113 |
| 494 | health promotion | 11 | 151 |
| 569 | information processing | 6 | 61 |
| 676 | mental health | 15 | 128 |
| 703 | mobile application | 6 | 67 |
| 707 | mobile phone | 6 | 20 |
| 759 | occupational diseases | 7 | 81 |
| 760 | occupational exposure | 12 | 142 |
| 762 | occupational health | 36 | 339 |
| 763 | occupational health and safety | 9 | 36 |
| 775 | occupational stress | 8 | 106 |
| 816 | pandemic | 12 | 159 |
| 817 | pandemics | 9 | 132 |

1. Coronavirus Disease 2019: With 10 occurrences and a total link strength of 151, this keyword reflects the significant impact of the COVID-19 pandemic on both digital health and occupational health domains. It highlights the heightened attention and research focus on addressing health challenges posed by the pandemic.
2. Depression: Appearing 11 times with a total link strength of 134, depression emerges as a prominent keyword, indicating the relevance of mental health issues within the context of digital and occupational health research. It underscores the importance of addressing psychological well-being in workplace environments.
3. Digital Technology: This keyword occurs 13 times and exhibits a total link strength of 148, emphasizing the pervasive influence of digital technologies in shaping contemporary approaches to healthcare delivery and workplace interventions. It highlights the increasing integration of digital solutions in occupational health practices.
4. Employee: With 8 occurrences and a total link strength of 113, the keyword “employee” underscores the central role of workforce engagement and management within discussions surrounding digital and occupational health. It reflects the focus on employee well-being and productivity enhancement strategies.
5. Health Promotion: Appearing 11 times and exhibiting a total link strength of 151, “health promotion” signifies the emphasis on proactive measures aimed at enhancing health outcomes and preventing illness within workplace settings. It underscores the importance of holistic approaches to occupational health management.
6. Mental Health: This keyword emerges prominently with 15 occurrences and a total link strength of 128, highlighting the growing recognition of mental health issues as significant factors influencing occupational well-being. It underscores the need for comprehensive mental health support in digital health interventions.
7. Occupational Health: With a staggering 36 occurrences and a total link strength of 339, “occupational health” emerges as a central theme in the dataset. It reflects the overarching focus of research endeavors on understanding and addressing health-related challenges within occupational settings.
8. Pandemic: Occurring 12 times and exhibiting a total link strength of 159, the keyword “pandemic” underscores the profound impact of global health crises on digital and occupational health landscapes. It emphasizes the urgency of responsive strategies and interventions to mitigate pandemic-related risks in workplace environments.

## 4 Discussion

The descriptive analysis of the research outcomes offers significant insights into the trends and dynamics of digital occupational health research. Firstly, the growth of publications over the years, as depicted in Figure 3, highlights a notable surge in research activity, particularly from 2017 onwards, with a peak in 2021. While there were fluctuations in annual publication counts, the cumulative trajectory demonstrates a consistent upward trend, indicating the progressive expansion of the scholarly discourse in this field.

The analysis of journal outlets, presented in Table 1, reveals the diverse landscape of publications contributing to digital and occupational health research. Notably, journals such as the International Journal of Environmental Research and Public Health, JMIR Mental Health, and PLOS ONE emerge as prolific platforms for disseminating research findings, with significant citation impact. The diversity of publishers and countries of origin underscores the global scope and interdisciplinary nature of research in this domain.

The examination of productive authors, countries, and institutions provides valuable insights into the collaborative networks and contributors shaping the scholarly landscape. Noteworthy contributions from authors such as Blake et al. and institutions like the University of Nottingham highlight the collaborative efforts driving advancements in digital occupational health research.

Furthermore, the subject area analysis, delineated in Table 2, elucidates evolving research priorities and interdisciplinary collaborations within the domains of digital and occupational health. The shift in subject areas over time reflects changing research emphases and emerging trends, such as the increased focus on environmental science and engineering in recent years.

Lastly, the bibliometric analysis of keywords offers valuable insights into thematic trends and focal points within digital occupational health research. The prominence of keywords such as “coronavirus disease 2019,” “depression,” and “digital technology” underscores the intersectionality of health crises, mental health concerns, and technological advancements in shaping research agendas. These findings collectively underscore the multifaceted nature of digital occupational health research and provide valuable guidance for future research endeavors and policy initiatives aimed at enhancing workplace health and well-being.

In recent years, there has been a notable shift towards exploring employee behaviors concerning occupational health, encompassing various dimensions such as medical consultations and conventional monitoring practices. The integration of digital technology has revolutionized occupational health practices, offering novel avenues for professionals to utilize tools like mobile applications, software, and E-consultations. These advancements enable practitioners to comprehensively document, analyze, and understand the intricate dynamics within workplace environments. Our study adhered meticulously to the PRISMA framework, systematically identifying relevant articles at the intersection of digital technology and occupational health. In essence, our research provides valuable insights into the evolving relationship between digital technology and occupational health, highlights key publishers and journals driving scholarly discourse, and underscores the impact of published literature in this rapidly evolving domain.

## Conclusion

Amidst the challenges posed by the Covid-19 pandemic, the significance of occupational health in safeguarding employee well-being and curbing the virus’s spread within the business environment has become increasingly evident. The integration of technology into occupational health practices has emerged as a vital strategy to reduce physical contact among employees and mitigate transmission risks. This study endeavors to illuminate the key themes and concepts encapsulated within the intersection of digital technology and occupational health through a rigorous bibliometric analysis.

The exploration of digital and occupational health themes has gained traction in recent years, particularly since 2019, reflecting a growing awareness of the interplay between technology and employee well-being. This surge in research activity is indicative of intensified efforts to comprehend and address health promotion strategies leveraging digital tools such as mobile applications and software. Over time, there has been a notable escalation in the volume of publications in this domain, witnessing a remarkable increase from just two articles in 2010 to over 20 articles in 2022.

This article provides a comprehensive examination of global trends in digital and occupational health research, identifying key countries, institutions, authors, journals, and application areas within the field. Analysis of the collected data enables quantification of the number of articles and citations, offering valuable insights into the scholarly impact of the selected literature. Noteworthy findings include the prominence of authors like Blake, H., the leadership of the United States in publication output, and the notable contributions of institutions such as the University of Nottingham and the University of St. Gallen.

While this study endeavors to address methodological limitations, including language bias towards English publications and a focus on technical and review papers, it acknowledges the presence of inherent biases and potential areas for improvement. Future research endeavors should strive to incorporate contributions from emerging nations and employ robust methodologies to ensure the reliability and validity of outcomes. In essence, this research provides a comprehensive overview of the evolving landscape of digital and occupational health, offering valuable insights into current trends and avenues for future exploration.

## Data Availability

All data produced are available online at Scopus

## References

1. Carolan, S., & De Visser, R. O. (2018). Employees’ perspectives on the facilitators and barriers to engaging with digital mental health interventions in the workplace: Qualitative study. JMIR Mental Health, 5(1) doi:10.2196/mental.9146

2. Blake, H., Lai, B., Coman, E., Houdmont, J., & Griffiths, A. (2019). Move-it: A cluster-randomised digital worksite exercise intervention in china: Outcome and process evaluation. International Journal of Environmental Research and Public Health, 16(18) doi:10.3390/ijerph16183451

3. Blake, H., Somerset, S., & Evans, C. (2020). Development and fidelity testing of the test@work digital toolkit for employers on workplace health checks and opt-in HIV testing. International Journal of Environmental Research and Public Health, 17(1) doi:10.3390/ijerph17010379

4. Blake, H., Somerset, S., & Greaves, S. (2022). The pain at work toolkit for employees with chronic or persistent pain: A collaborative-participatory study. Healthcare (Switzerland), 10(1) doi:10.3390/healthcare10010056

5. Block, J. H., & Fisch, C. (2020). Eight tips and questions for your bibliographic study in business and management research. Management Review Quarterly, 70(3), 307–312. DOI:10.1007/s11301-020-00188-4

6. Carolan, S., Harris, P. R., Greenwood, K., & Cavanagh, K. (2017). Increasing engagement with an occupational digital stress management program through the use of an online facilitated discussion group: Results of a pilot randomised controlled trial. Internet Interventions, 10, 1–11. doi:10.1016/j.invent.2017.08.001

7. Halldin, C. N., Hale, J. M., Weissman, D. N., Attfield, M. D., Parker, J. E., Petsonk, E. L.,. Laney, A. S. (2019). The national institute for occupational safety and health B reader certification program - an update report (1987 to 2018) and future directions. Journal of Occupational and Environmental Medicine, 61(12), 1045–1051. doi:10.1097/JOM.0000000000001735

8. Judson, T. J., Odisho, A. Y., Young, J. J., Bigazzi, O., Steuer, D., Gonzales, R., & Neinstein, A. B. (2020). Implementation of a digital chatbot to screen health system employees during the COVID-19 pandemic. Journal of the American Medical Informatics Association, 27(9), 1450–1455. doi:10.1093/jamia/ocaa130

9. Moher, D., Shamseer, L., Clarke, M., Ghersi, D., Liberati, A., Petticrew, M., … Stewart, L. A. (2015). Preferred reporting items for systematic review and meta-analysis protocols (PRISMA-P) 2015 statement. Systematic reviews, 4(1), 1–9. DOI:10.1186/2046-4053-4-1

10. Van Eck, N., & Waltman, L. (2013). Manual for VOSviewer version 1.5. 4. Universiteit Leiden and Erasmus Universiteit Rotterdam, 1(1), 1–53.

11. Wang, M., & Chai, L. (2018). Three new bibliometric indicators/approaches derived from keyword analysis. Scientometrics, 116(3), 721–750. DOI:10.1007/s11192-018-2768-9

12. Zhang, Y., Wu, X., Gao, J., Chen, J., & Xv, X. (2019). Simulation and ergonomic evaluation of welders’ standing posture using jack software. International Journal of Environmental Research and Public Health, 16(22) doi:10.3390/ijerph16224354.

13. Er Rays Youssef, Hamid Ait Lequaddem, et Mustapha Ezzahiri. 2024. « Bibliometric Analysis of Global Research Trends on Digital Occupational Health Using Scopus Database ». P. 1-12 in International Conference on Advanced Intelligent Systems for Sustainable Development (AI2SD’2023), édité par M. Ezziyyani, J. Kacprzyk, et V. E. Balas. Cham: Springer Nature Switzerland.

14. Er-Rays, Youssef, et Hamid Ait-Lemqeddem. 2020. « La performance hospitalière au Maroc et COVID-19 : Application d’Analyse d’Enveloppement des Données et l’indice de Malmquist ». International Journal of Accounting, Finance, Auditing, Management and Economics 1(2):334–52. doi: 10.5281/zenodo.4027715.

15. Er-Rays, Youssef, et Hamid Ait Lemqeddem. 2020. « La performance des établissements des soins de santé de bases au Maroc et COVID-19 : Application d’Analyse d’Enveloppement des Données et l’indice de Malmquist ». Revue Française d’Economie et de Gestion 1(4).

16. Er-Rays, Youssef, et Hamid Ait Lemqeddem. 2021. « La performance de Système de Santé Marocain et COVID-19 entre le système bismarckien et le système beveridgien ». https://ijarims.org/volume-4-issue-1-html/# 4(1):17.

17. Er-Rays, Youssef, et Meriem M’dioud. 2024a. « Assessment of Technical Efficiency in the Moroccan Public Hospital Network: Using the DEA Method ».

18. Er-Rays, Youssef, et Meriem M’dioud. 2024b. « Evaluating the Financial Factors Influencing Maternal, Newborn, and Child Health in Africa ».

19. Er-Rays, Youssef, Meriem M’dioud, Hamid Ait-Lemqedde, et Mustapha Ezzahir. 2024a. « Data Envelopment Analysis and Malmquist Index Application: Efficiency of Hospitals Networks in Morocco ». P. 13-24 in International Conference on Advanced Intelligent Systems for Sustainable Development (AI2SD’2023). Vol. 904, Lecture Notes in Networks and Systems, édité par M. Ezziyyani, J. Kacprzyk, et V. E. Balas. Cham: Springer Nature Switzerland.

20. Er-Rays, Youssef, Meriem M’dioud, Hamid Ait-Lemqedde, et Mustapha Ezzahir. 2024b. « Data Envelopment Analysis and Malmquist Index Application: Efficiency of Hospitals Networks in Morocco ». P. 13-24 in International Conference on Advanced Intelligent Systems for Sustainable Development (AI2SD’2023), édité par M. Ezziyyani, J. Kacprzyk, et V. E. Balas. Cham: Springer Nature Switzerland.

